# Effects of Parkinson’s Disease on Mechanical and Microstructural Properties of the Brain

**DOI:** 10.1101/2025.03.24.25324514

**Authors:** Christoffer Olsson, Mikael Skorpil, Per Svenningsson, Rodrigo Moreno

## Abstract

Magnetic Resonance Elastography (MRE) is a method capable of mapping the brain’s mechanical properties, however, the microstructural mechanisms responsible for these biomechanical properties remain largely unknown. For this reason, the present study utilized multidimensional diffusion-MRI (MD-dMRI), apart from MRE, to extract microstructural parameters for a cohort of Parkinson’s disease (PD) patients and healthy controls. Significant softening effects in the temporal and occipital lobes in PD were associated with an increase in mean diffusivity, whereas other microstructural properties, e.g. microscopic FA (µFA), largely remained unchanged. Across most regions, stiffness declined with age, which was correlated with a decrease in μFA and an increase in MD. We hypothesize that age softening effects mostly can be explained by neuronal atrophy, whereas PD effects involve additional mechanisms.

## 1. Introduction

Parkinson’s disease (PD) is a neurodegenerative disease that affects about 1% of the population over the age of 60^1,2^. It is characterized by a progressive loss of dopaminergic neurons in the substantia nigra, which decreases the patient’s dopamine levels leading to motor function impairments, such as bradykinesia, ridigity and tremor. Apart from motor functions, PD patients typically also exhibit other symptoms, such as cognitive decline and depression. Traditional MRI techniques have revealed substantial macrostructural changes in various brain regions due to atrophy in PD^3^, especially in the advanced stages of the disease. Less conventional techniques, such as quantitative susceptibility mapping (QSM) and neuromelanin imaging (NMI), have also been used to depict differences between PD and healthy controls^4–7^ by quantifying the amount of iron and neuromelanin, respectively, in various areas of the brain. Any alterations due to PD in macroscopic measures arise as a consequence of microstructural changes in the brain. Thus, assessing microstructural changes in PD might give hints of the mechanisms of the disease that lead to macrostructural changes, such as volumetric reduction of brain tissue, iron, or neuromelanin content.

One method of obtaining microstructural information is multidimensional diffusion magnetic resonance imaging (MD-dMRI), which is an advanced diffusion sequence that combines both a multi-shell diffusion MRI (dMRI) acquisition and a spherical b-tensor encoding to obtain different microstructural parameters, such as microscopic anisotropy or free water fractions and has previously been used to investigate PD^8–10^. The main advantage of this MRI modality is that it can distinguish between different structural configurations at the microscale that are impossible to detect with standard processing of dMRI data.

Furthermore, the mechanical properties (e.g. stiffness and viscosity) of the brain can provide additional insights into the brain’s microstructure, thus investigations on how these properties change due to PD may provide valuable information about the disease. For example, studies on Alzheimer’s disease, have shown that changes in the mechanical properties, as measured by magnetic resonance elastography (MRE)^11^, can appear before they are visible in structural MRI^12^. MRE is a technique that can estimate the mechanical properties of the brain in vivo, in particular, shear stiffness and viscosity. For MRE, the subject’s head is gently vibrated inside of an MR scanner, while a set of motion encoding gradients are employed to detect small displacements of the tissue from the phase of the MR signal. A map of the mechanical properties of the brain can then be reconstructed based on these displacement images. MRE has been used to study several different aspects of the brain and is emerging as an increasingly promising imaging modality^13^. For example, studies have found that the brain generally softens with increasing age^14,15^. Moreover, it has been found that the mechanical properties of the brain are affected by pathologies like Alzheimer’s disease^12,16^, multiple sclerosis^17,18^, and PD^19,20^.

The mechanical properties of the brain measured by MRE at a voxel level are determined by the microstructural properties of the brain, i.e., by its structural composition of neurons, glial cells, blood vessels, and extracellular water, among others. Extensive efforts have been made to study how the microstructure relates to the stiffness of healthy and diseased brain tissue^21–24^. Still, the understanding of microstructure and mechanics of the brain *in vivo* is limited due to few studies utilizing imaging techniques that extract such properties.

This study contributes to closing the gap between macro and microstructural information by assessing the relationships between the mechanical properties estimated with MRE and microstructural parameters estimated with multidimensional diffusion MRI (MD-dMRI) in PD. Apart from these, we also used quantitative susceptibility mapping (QSM) and neuromelanin imaging (NMI). To the best of our knowledge, this is the first study where MD-dMRI has been employed to provide a microstructural explanation for macroscopical property changes measured with MRE, QSM, and NMI on PD.

## 2. Methods

### 2.1. Subject information

A description of the population is shown in Table 1. The cohort consists of 17 healthy controls (HC) and 12 PD subjects in ON state. The disease severity of the PD subjects was evaluated by a clinician using the Unified Parkinson’s Disease Rating Scale (UPDRS I-IV). We obtained a written informed consent form prior to the participation from all subjects and obtained ethical approval from the Swedish Ethical Review Authority (Dnr. 2022-03209-02).

**Table 1.** Subject information.

| Subject Info | Controls | PD Patients |
| --- | --- | --- |
| Mean age ± SD [min-max] | 60 ± 6 [50-78] | 63 ± 9 [47-78] |
| Gender [F:M] | 3:14 | 3:9 |
| Disease Duration [Years] | -- | 6.4 ± 5.3 [1-17] |
| LEDD [mg] | -- | 479 ± 301 [72-958] |
| H & Y | -- | 1.75 ± 0.45 [1-2] |
| MoCA | -- | 28.9 ± 1.2 [27-30] |
| MDS-UPDRS-1 | -- | 7.0 ± 2.3 [2-10] |
| MDS-UPDRS-2 | -- | 6.5 ± 2.3 [3-11] |
| MDS-UPDRS-3 | -- | 18.7 ± 11.0 [4-41] |
| MDS-UPDRS-4 | -- | 5.1 ± 4.1 [0-12] |
| Total MDS-UPDRS | -- | 35.3 ± 9.8 [24-58] |

### 2.2. Image acquisition

All subjects were scanned using a Philips Ingenia CX 3T scanner with a 32-channel head coil. Participants’ heads were placed on top of a pneumatically driven vibrating pillow that was used for the MRE scan at the end of the session. The session included standard T1-weighted and T2-weighted images. T1w: Turbo Field Echo sequence, TR/TE: 6.7/3 ms, 1×1×1mm^3^ resolution, full head FOV, T2w: TR/TE: 3000/280 ms, 1×1×1mm^3^ full head FOV. MD-dMRI images were acquired as explained in Ref.^25^ with TR/TE: 4000/111 ms and a resolution of 2.5×2.5×2.5 mm^3^, using spherical and linear b-tensors with 5 b-values (b=0,100,700,1400,2000s/mm^2^). The MD-dMRI acquisition lasted for a total of 6.5 minutes. MRE images were acquired using a spin-echo EPI sequence with 3-directional motion encoding gradients, with a gradient strength of 70mT/m, TR/TE: 4800/67 ms, 3×3×3mm^3^ with full brain FOV^*^. The driver pillow was set to vibrate with a driving frequency of 60Hz and was provided by Mayo Clinic (see, e.g., Ref. ^26^ for more details on the acquisition protocol). The acquisition lasted for about 4.5 minutes.

To obtain QSM images, 8 different gradient-recalled-echo (GRE) images with different echo times linearly spaced between 9.8 ms-57.4ms were acquired with a TR of 62ms, a flip angle of 15° and a spatial resolution of 1×1×1 mm^3^.

NMI was obtained with a gradient echo sequence with a FOV of 210x210x46.8mm^3^ with 36 1.3mm thick slices centered at the midbrain and in-plane resolution of 0.6 mm, TR/TE: 83/7.7 ms and a flip angle of 16°. The acquisition lasted for about 5.25 minutes. The obtained NMI-intensity was finally normalized to 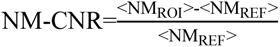 where <NM_ROI_> is the mean intensity within the region of interest, and <NM_REF_> is the mean intensity within a reference region typically containing little neuromelanin, in our case we chose the WM-temporal as reference, in line with Bashat et al^27^.

Each scanning session lasted about 45 minutes and included other imaging modalities not relevant to the work of this paper, all acquired before the MRE scan.

### 2.3. Postprocessing

#### MRE

Regarding MRE, the acquired displacement images were inverted to viscoelastic parameter maps as described in Ref. ^28^. This results in an image with the complex shear modulus per voxel, which is defined as G*=G’+iG’’. Stiffness is defined as |G*|, and the viscosity-related phase angle φ = arctan(G′′/G′).

#### MD-dMRI

Regarding MD-dMRI, Synb0-DISCO^29,30^ was used to produce synthetic distortion correction images for the MD-dMRI images for subsequent topup correction using FSL ^31,32^. The images were post-processed using the open-source MD-dMRI software^33^ to estimate four parameters: fractional diffusivity (FA), mean diffusivity (MD), microscopic FA (μFA) and the normalized variance of MD (here referred to as Var(MD), see parameter CMD in Ref. ^34^). From these, μFA and Var(MD) cannot be estimated with standard diffusion MRI (dMRI). These parameters were estimated using methods described in Westin et al^34^. Figure 1a sketches the difference between FA and μFA. FA is conventionally obtained from standard dMRI; however, this parameter is a macroscopic average measure of anisotropy; thus, if a volume contains, e.g., axons with different orientations, the average anisotropy will be very low. In contrast, μFA measures microscopic anisotropy; thus, the same volume would result in a high μFA. If the volume has a high density of aligned fibers, both FA and μFA are high. In turn, if the voxel exhibits high MD (see Figure 1b), this means that the volume contains a lot of freely diffusing water molecules, and if there is a low Var(MD), the obtained MD signal is drawn from a homogeneous distribution of water molecules with similar diffusivity, however, if the Var(MD) is high this distribution is inhomogeneous; the latter means that there are several different populations of water molecules within the volume (e.g., intra- and extracellular water) that diffuses with different rates.

**Figure 1.**
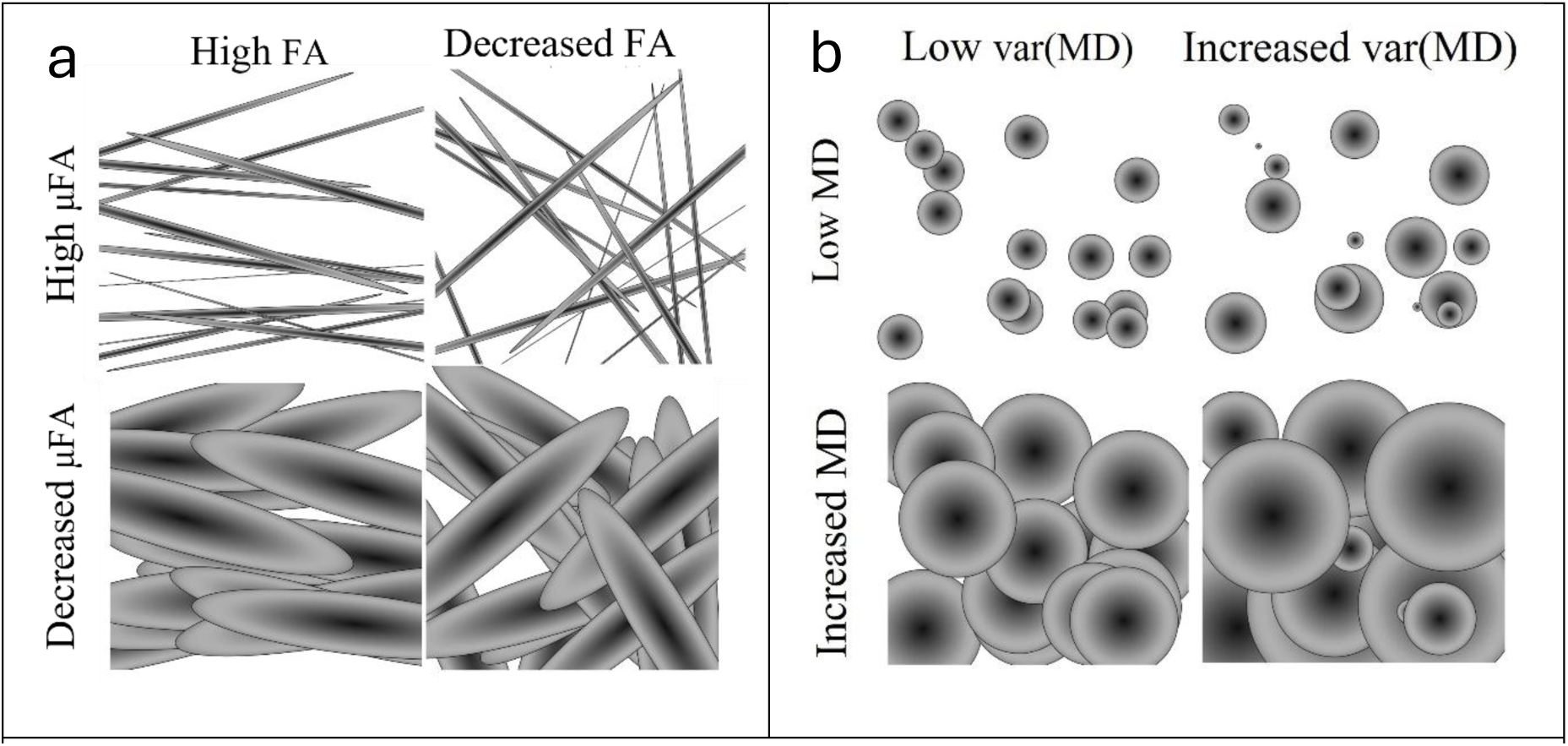
Schematic for how subvoxel properties change with varying MD-dMRI parameters: a) FA vs.μFA and b) MD vs. Var(MD) for different sub-voxel scenarios. a) high µFA means that there are plenty of water compartments that diffuses with high anisotropy, and by altering the alignment of these compartments one can obtain either high or low average anisotropy (FA). Decreasing µFA implies either that there are fewer compartments of high anisotropy, or that the anisotropy of the compartments decreases. b) MD measures the mean diffusivity of the entire voxel and the Var(MD) is a measure for the normalized variance of MD for the different subpopulations within the voxel.

#### QSM

The images acquired with the QSM protocol were postprocessed with the use of the morphology-enabled dipole inversion (MEDI)-toolbox ^35^ to invert the phase images and obtain the final QSM.

#### Spatial normalization

All parameters extracted from MRE, MD-dMRI, QSM and NMI were, after postprocessing, registered to subject-specific T1 images (1×1×1mm^3^), which had been parcellated into the Desikan-Killiany atlas ^36^ with the use of FreeSurfer v7.2. Values from substructures in the brainstem (mesencephalon and pons) were also extracted using an additional segmentation method within FreeSurfer ^37^. These regions were then combined into various groups (e.g., the cortical layer of the occipital lobe). Median values of each of these regions were then extracted for statistical analysis.

To analyze subcortical structures we utilized a high-resolution atlas (in MNI space) of the following structures^38^:

- Putamen (Pu)
- Caudate Nucleus (Ca)
- Nucleus Accumbens (NAC)
- Globus Pallidus external and internal (GPe and GPi)
- Substantia Nigra pars compacta and pars reticulata (SNc and SNr)
- Red Nucleus (RN)
- Ventral Tegmental Area (VTA)

The subject-specific T1w images were first registered to an MNI template using affine registration, followed by a nonlinear registration with the use of ANTs ^39^. The inverses of these registrations were then applied to the subcortical atlas to obtain the subject-specific parcellation of the subcortical structures in the subject space.

## 3. Results

### 3.1. MR Elastography

Figure 2a shows the brain stiffness averaged for the HC and PD subjects after registering the individual maps to MNI space^40^. The stiffness for subjects with PD can be seen generally to be lower than for the HC. Figure 2b shows the average stiffness and 2c shows average viscosity (averaged over the entire brain, excluding ventricles and CSF) as a function of the individual subject’s age for both HC (marked in black) and PD (marked in red). There, it becomes apparent that the brain’s stiffness decreases with age, however, the effect is more prominent in PD subjects. Viscosity also exhibits a decreasing trend with age, however the difference between HC and PD is less obvious.

**Figure 2.**
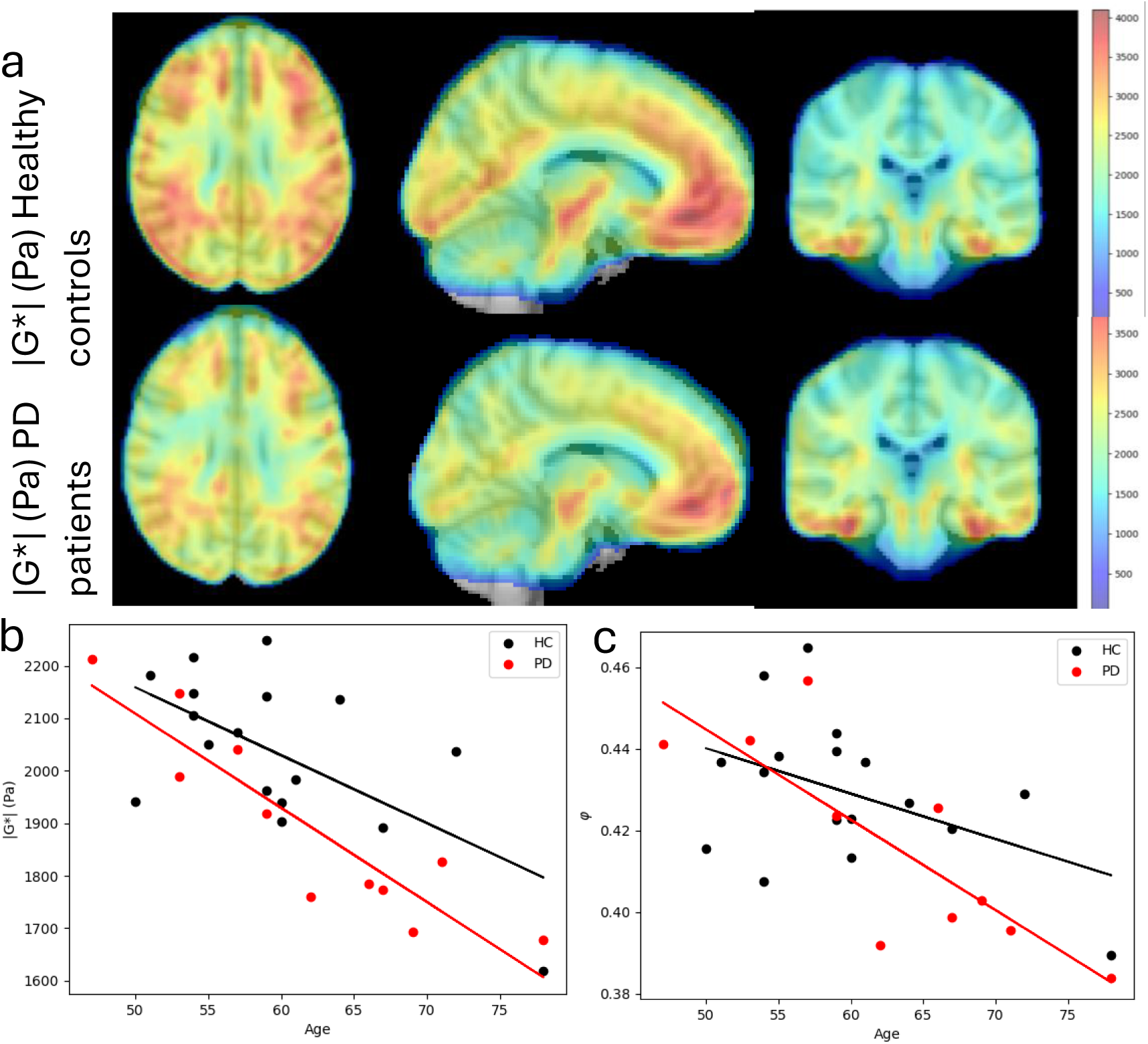
a. Brain stiffness averaged for all subjects in MNI space. Top: healthy controls. Bottom: PD patients. Columns show axial, sagittal, and coronal views, respectively. The scale bar shows stiffness values given in Pa. 2b Average whole brain stiffness |G*| and 2c viscosity angle *φ* (in radians) vs subject age for healthy controls (black points) and patients with PD (red points). Regions containing CSF were excluded from the average.

### 3.2. MD-dMRI

Figure 3a shows average μFA-maps for all HC and PD subjects, where some small local differences can be seen between the two groups. For example, there is slightly higher μFA in the frontal and temporal lobes for HC than for PD. Figure 3b-e shows how the various MD-dMRI-derived parameters change, on average, across the whole brain as a function of age. It is clear that MD (and, to a lesser degree, Var(MD)) exhibits a positive correlation with age, whereas μFA exhibits a negative correlation with age. No significant correlation was detected for FA. No significant differences can be seen between HC and PD for the whole brain averages of the MD-dMRI parameters.

**Figure 3.**
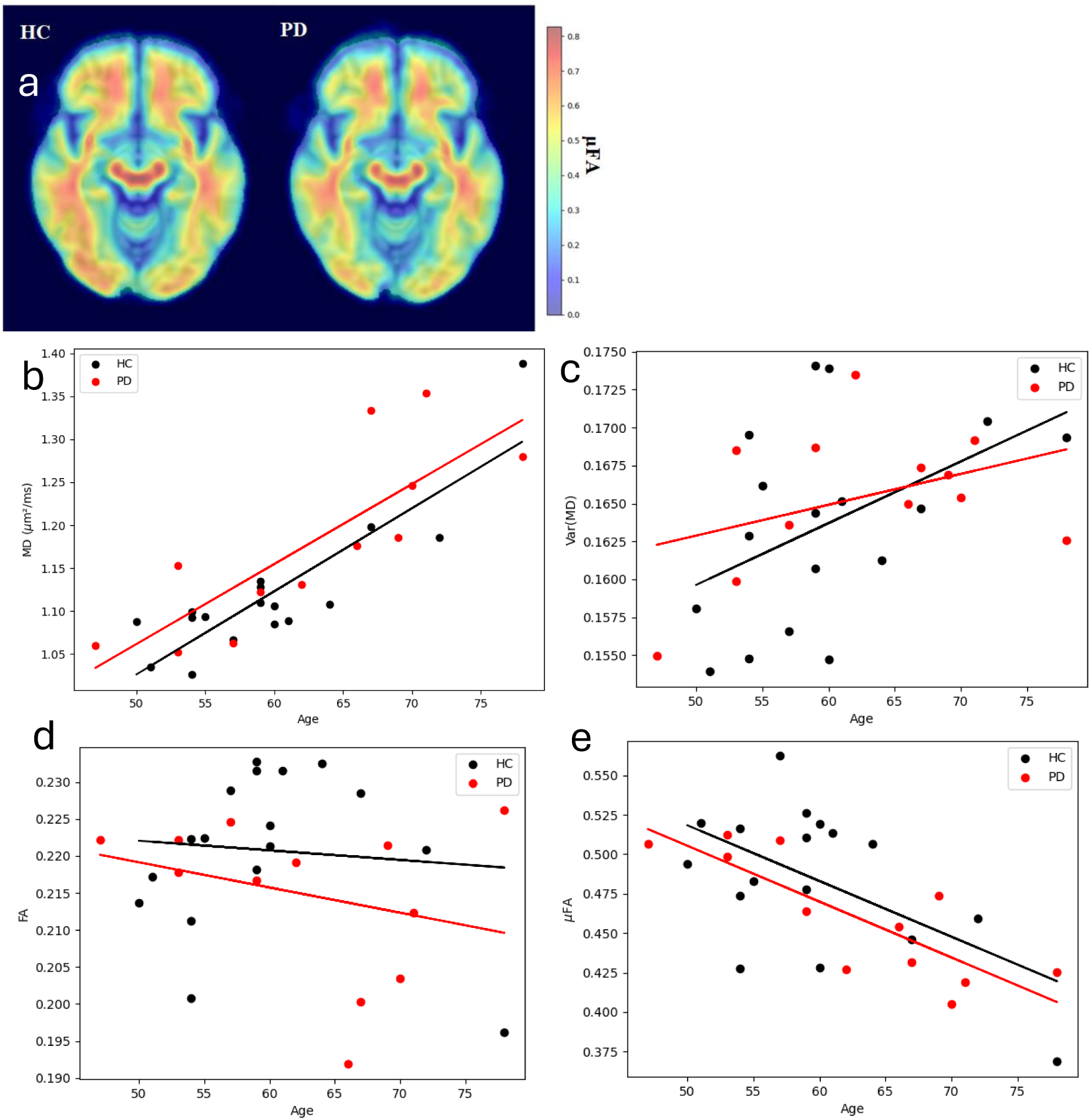
a) MD-dMRI parameter example maps for μFA averaged over all healthy controls and all PD patients registered to the MNI space. b) – e) -MD-dMRI parameters averaged over the whole brain as a function of age. Regions containing CSF are excluded. Black and red points indicate healthy controls and PD patients, respectively. Linear fits are shown as a general trend line for the different groups.

### 3.3. Age effects

Figure 4a shows the correlations between MD-dMRI and MRE, QSM and NMI measurements with the age of the subjects subdivided into various brain regions. The brain regions were subdivided into the cortex (Ctx) and white matter (WM) of the four main lobes of the cerebrum (frontal, parietal, temporal, and occipital), and additionally, the corpus callosum and the mesencephalon. The correlations were calculated based on all subjects and corrected for group (PD or HC) as a covariate.

**Figure 4.**
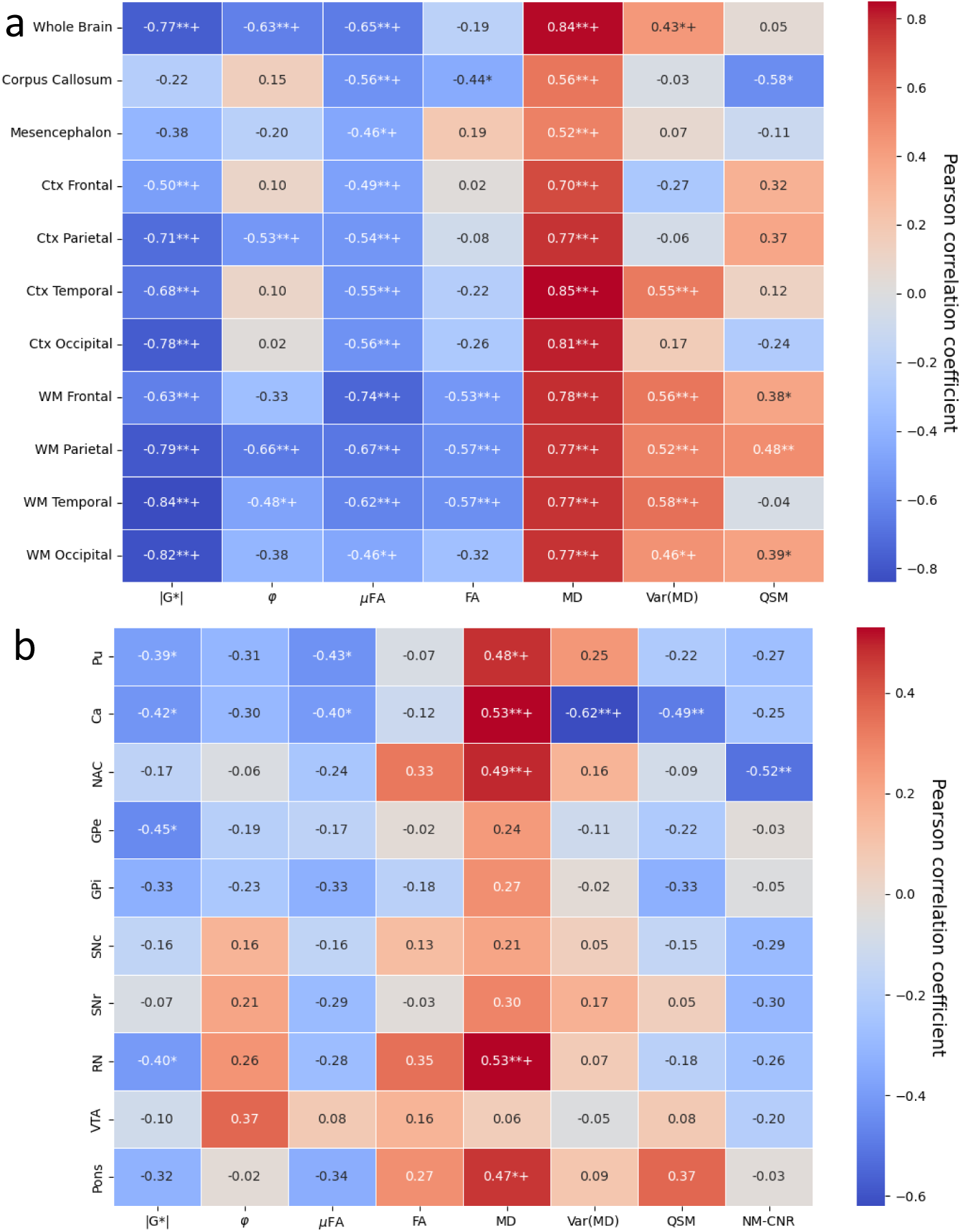
Age effects. Pearson correlations between age and mean intensity of a) major brain regions and b) subcortical regions, corrected for group effects (PD or HC). (*=p<.05. **=p<0.01. += p_FDR-corr <0.05). Pu=Putamen, Ca=Caudate Nucleus, NAC=Nucleus Accumbens, GPe/i =Globus Pallidus external/internal, SNc/r=Substantia Nigra pars compacta/reticulata, RN=Red Nucleus, VTA=Ventral Tegmental Area

Apart from the corpus callosum and the mesencephalon, all structures appear to soften significantly with age (after false discovery rate Benjamini-Hochberg (FDR-BH) correction) with varying degrees, where the white matter regions of the cerebrum are the most affected. Fewer significant effects can be seen on the viscosity (φ), but still, the parietal lobe, the WM temporal, and the whole brain in general can be seen to significantly decrease in viscosity.

The μFA is significantly decreased for all structures due to age, with the WM of the frontal lobe most significantly affected. This effect is strongly correlated with increased MD, which is significantly increased in all regions. The normalized variance of MD (Var(MD)) is most significantly shown to correlate positively with age for the WM of the cerebrum and on the Ctx of the temporal lobe, with little effect on the remaining structures. The macroscopic fractional anisotropy (FA) is most significantly decreased in the parietal, frontal and temporal WM, and to a lesser extent in the CC, with little effect on other structures. Note that both μFA and FA are reduced for higher age for structures containing large WM tracts (cerebrum WM, corpus callosum, and mesencephalon), whereas in the Ctx, FA is not significantly dependent on age (FA values in the Ctx are very low in general, as expected).

Within the subcortical structures (see Fig. 4b), there is a significant softening (reduced |G*|) of the Putamen (Pu), Caudate nucleus (Ca), Globus pallidus exterior (GPe), and Red Nucleus (RN), with p<0.05, p_FDR-corr>0.05. No significant effects were seen for the viscosity parameter. Some significant results were found for the μFA on the Pu, and Ca (p<0.05, p_FDR-corr>0.05), but not for the FA. MD, however, displays several significant positive correlations with age within the Pu, Ca, NAC, RN and pons (p_FDR-corr<0.05 for all). Var(MD) only shows a significant decrease with age within the Ca (p<0.01, p_FDR-corr<0.05).

The effect of gender on the various modality measures was not taken into account due to the small sample size (only three women in either group).

The tissue volume for several larger regions was compared for HC and PD patients, and although there was a general trend for smaller volumes in PD compared to HC, these differences were non-significant (two-sided t-test p>0.05 for all comparisons).

### 3.4. Effects of Parkinson’s disease

Figure 5 shows correlations (effect sizes, calculated as Cohen’s d between the two groups, and the associated p-values were calculated based on the Mann-Whitney U test) between the median imaging modality values in the different selected regions of the brain after correcting for age effects (age effect correction was applied by adjusting the data due to age by linear regression of the data vs. age and subtracting this effect from the data). Group comparisons with unadjusted values are shown in the SI (Figs. SI6 and SI7). On average, over the whole brain, the only statistically significant change found was a decrease in stiffness (p<0.05, p_FDR-corr>0.05). Zooming in on the different regions, we found a softening of the temporal lobe (Ctx: p<0.05, p_FDR-corr>0.05; WM: p<0.05, p_FDR- corr>0.05), the parietal Ctx (p<0.05 p_FDR-corr>0.05) and for the occipital Ctx ( p<0.01, p_FDR-corr>0.05). Although not always significant (p>0.05), the effect sizes show that a decrease in stiffness is positively correlated with a decrease in μFA and negatively correlates with increasing MD and increasing Var(MD) for the temporal and occipital lobes.

**Figure 5.**
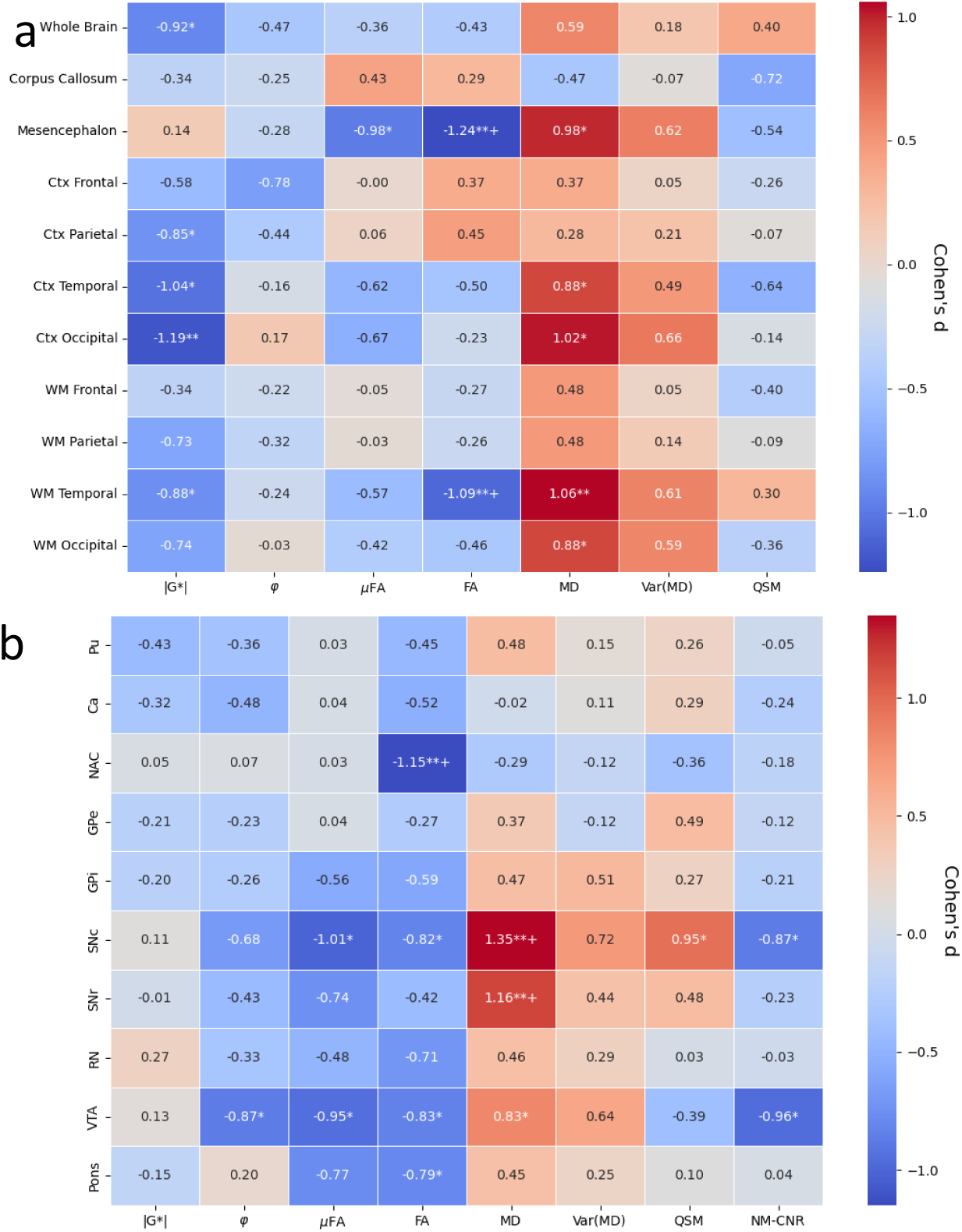
Standardized effect sizes (Cohen’s d) for the two groups (PD or HC). Median values in ROIs were adjusted for age effects by linear regression. (p values calculated by Mann-Whitney U-test between groups with adjusted values, *=p<.05. **=p<0.01. += p_FDR-corr <0.05). a) shows major brain regions, and b) shows minor subcortical regions. Pu=Putamen, Ca=Caudate Nucleus, NAC=Nucleus Accumbens, GPe/i =Globus Pallidus external/internal, SNc/r=Substantia Nigra pars compacta/reticulata, RN=Red Nucleus, VTA=Ventral Tegmental Area

The softening of these regions is also accompanied by an increase of MD (p<0.05 for temporal Ctx, p<0.01/p_FDR-corr<0.05 for occipital Ctx, p<0.01 for temporal WM, and p<0.05 for occipital WM). Interestingly, the mesencephalon, being strongly affected by PD, did not exhibit any significant softening in this study, as opposed to what was reported in previous studies^19^. The mesencephalon did however exhibit a reduction in µFA (p<0.05, p_FDR.corr>0.05) and FA (p<0.01, p_FDR.corr<0.05), and an increase of MD (p<0.05, P_FDR-corr>0.05).

For the subcortical regions (Figure 5b), we again see no significant softening due to PD, and little effect on viscosity in any region (with the exception of the VTA). There are, however, some significance for the μFA within the SNc (p<0.01 p_FDR-corr>0.05) and VTA (p<0.05 p_FDR- corr>0.05); some for FA within NAC (p<0.01 p_FDR-corr<0.05), and the SNc and the VTA (p<0.05 p_FDR-corr>0.05). Strong significance can be seen for group effects of MD (positive correlation) within the SNc, SNr, mesencephalon (p<0.01 p_FDR-corr<0.05) and VTA (p<0.05 p_FDR- corr>0.05). The Var(MD) follows the same pattern as the MD, but with very little to no significance.

### 3.5. QSM and NMI results

As seen in fig 4a, levels of iron, as indicated by QSM levels in the brain, are increasing with age in some regions, most significantly in the parietal (p<0.01, p_FDR-corr>0.05) frontal and occipital WM (p<0.05 p_FDR-corr>0.05), and decreasing with age in the corpus callosum (p<0.05 p_FDR- corr>0.05). Neuromelanin was excluded from the major brain region analysis since the FOV was limited to a slab crossing the midbrain, and thus most global structures, such as the occipital lobe, are only partially included and may vary between subjects. Within the subcortical structures (Fig 4b), QSM only shows a negative significant correlation with age within the Ca (p<0.01, p_FDR- corr>0.05), whereas NM-CNR shows significant negative correlations for the NAC (p<0.01, p_FDR- corr>0.05).

For group differences between PD and HC (see Fig 5a), there are no significant differences on the global level after FDR-corrections for QSM, although the QSM signal in the corpus callosum and Ctx temporal exhibits large effect size (|Cohen’s d|>0.6). Zooming in on the subcortical regions (Fig. 5b), however, we detect a significant increase of QSM in the SNc (p<0.05, p_FDR-corr>0.05), and a decrease of NM-CNR in the SNc, and the VTA ( p<0.05, p_FDR-corr>0.05).

### 3.6. Correlations between MRE, MD-dMRI and QSM

Correlations between all image modalities were calculated for the whole brain to assess the relationship between MRE, MD-dMRI and QSM parameters. Figure 6a shows these correlations for the whole brain averages. Figure 6b shows an example correlation plot for several regions overlaid, specifically for |G*| as a function of MD. Correlations between other imaging modalities can be found in the SI (SI Figs. 8-10). There exists a clear negative correlation between MD and both stiffness and viscosity and a clear positive correlation between μFA and both stiffness and viscosity. These correlations are well in line with the results from Ref. ^41^, who showed a significant negative correlation between the apparent diffusion constant and the stiffness of the liver. Notice also that the correlation between Var(MD) and stiffness is weak, and that QSM has little correlation to any of the other parameters.

**Figure 6.**
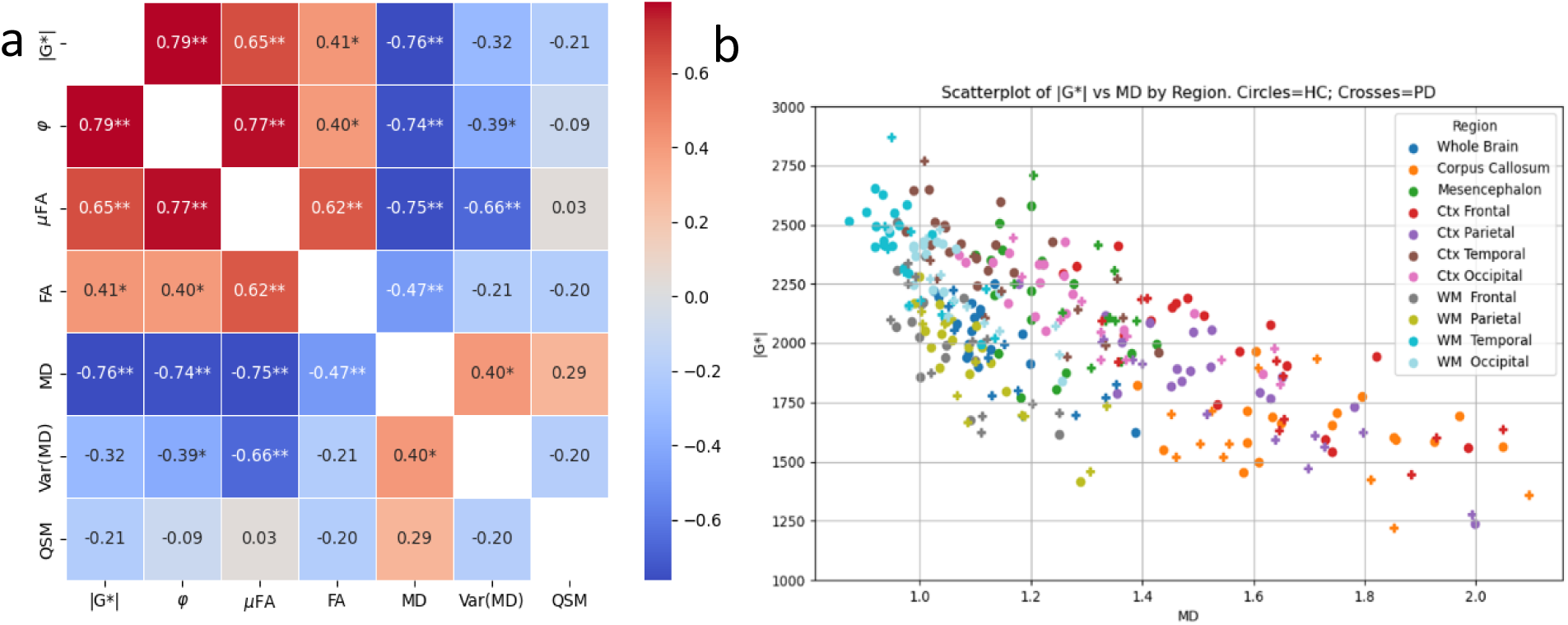
a) Correlations between imaging modalities for all subjects’ whole brain average. Values in each box show the Pearson correlation coefficient after correction for group (HC vs. PD) effects. * = p<0.05, ** = p<0.01. b) Stiffness as a function of MD for all subjects divided into different regions, as listed in the legend. Circular marks signify HC, plus-signs signify PD subjects.

### 3.7. Correlations within subcortical structures with clinical scores (UPDRS)

The various extracted measurements were also evaluated against the different clinical UPDRS-scores (UPDRS 1-4) of the PD patients. Pearson correlations between UPDRS scores and imaging modality parameters were calculated after correcting for age and are shown in the SI (Figs. SI3-SI5). Figure 7 shows the Pearson correlations between the various imaging modalities and the total UPDRS score (sum of UPDRS 1-4), where most of the correlations are relatively insignificant for the global regions (see Fig 7a), showing only a significant increase in μFA for the whole brain and corpus callosum (p<0.05, p_FDR-corr>0.05). Zooming in to the subcortical regions (see Fig. 7b), we find negative correlation between total UPDRS and |G*| within the pons (p<0.05, p_FDR-corr>0.05), and negative correlation between total UPDRS and QSM within the GPi (p<0.05, p_FDR-corr>0.05).

**Figure 7.**
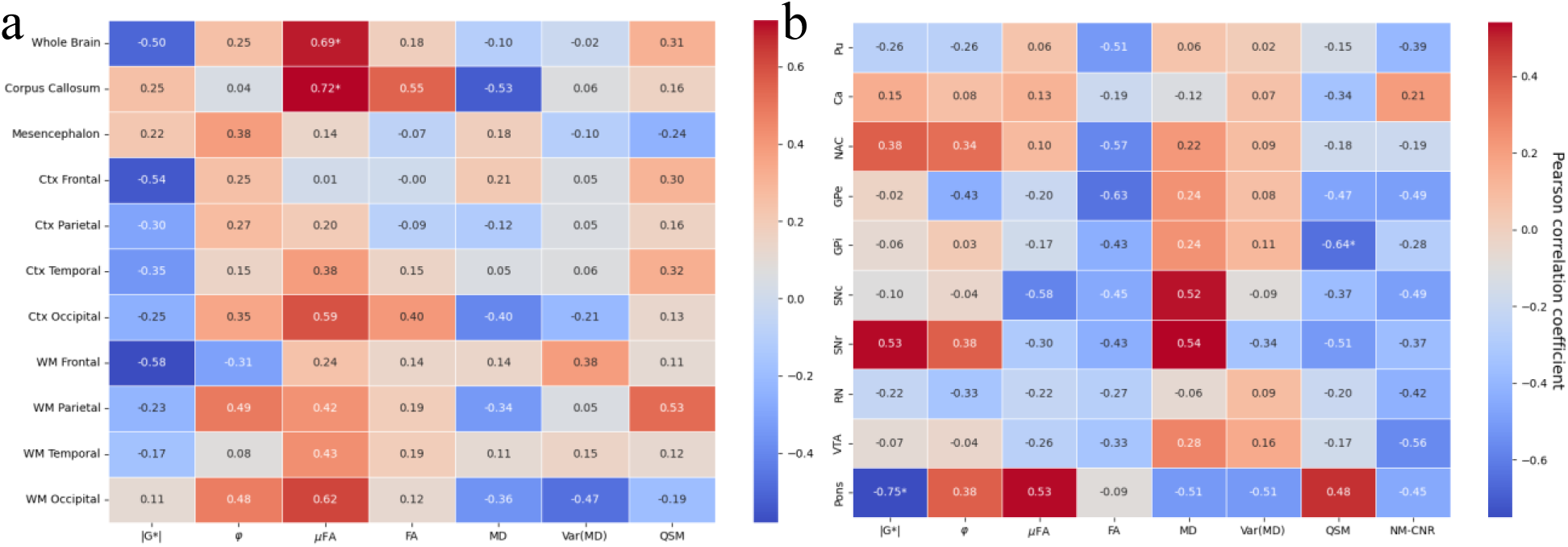
Correlations (Pearson coefficients) between changes in the measured values with total UPDRS score (age-corrected) for the PD cohort (excluding HC). a: global structures. b: subcortical structures. *=p<0.05, **=p<0.01, +=p_FDR-corr<0.05. Pu=Putamen, Ca=Caudate Nucleus, NAC=Nucleus Accumbens, GPe/i =Globus Pallidus external/internal, SNc/r=Substantia Nigra pars compacta/reticulata, RN=Red Nucleus, VTA=Ventral Tegmental Area

## 4. Discussion

The reduction of shear stiffness with age has previously been shown and, for example, Sack et al^15^ found that |G*| is reduced with 15 Pa per year for healthy subjects. This trend matches the present results (see Fig. 2b). For PD patients, on the other hand, |G*| is reduced by approximately 20Pa per year (see Fig. 2b), suggesting an accelerated aging effect.

Only a few studies have investigated the effects of PD on the mechanical properties of the brain using MRE in humans. In particular, two studies by Lipp et al ^19,20^ have investigated this relationship in two cohorts of 18 PD vs 16 HC in Lipp et al. (2013)^20^ and 17 PD vs 12 HC in Lipp et al. (2018)^19^, respectively^†^. Notice that those studies^19,20^ were carried out with different scanner settings, which may have a significant effect on the measured viscoelastic properties^42^. For example, they only acquired a part of the brain (6 cm slab through the central cerebrum in the 2013 study and a 2 cm slab in the 2018 study), and they used a different frequency (50Hz) in the 2013 paper and multifrequency acquisition (6 frequencies at 30-60Hz) in the 2018 paper, which makes it difficult to make quantitative comparisons. Another important work on PD using MRE is that of Hain et al^21^, who selectively removed dopaminergic neurons in the substantia nigra of mice (by injection of 1-methyl-4- phenyl-1,2,3,6-tetrahydropyridin hydrochloride (MPTP)) and measured the effect of the resulting viscoelastic parameters. They found that the effects of MPTP induced neurodegeneration of dopaminergic neurons in the SN, which correlated with decreased viscosity and decreased elasticity to a less significant extent.

As previously mentioned, our results suggest that the stiffness of the brain generally decreases with PD, which is in line with the findings of Lipp et al^19,20^ (comparisons between the present study with these two studies by Lipp et al. can be found in the supplementary information) and analogous to the findings in a mice model by Hain et al^21^. Although the measurements from Ref. ^19^ deviated from our results (and their previous study^20^), the relative changes due to PD are comparable. For example, Table SI2 shows that in the frontal region, we detect an 8% reduction (p=0.04) for the HC/PD group effect (using an ANOVA test with age as a covariate) compared to Lipp et al^19^, who reported a 14% decrease in |G*| (p=0.02) in the frontal lobe (although they used a different segmentation method).

Lipp et al^19^ also detected a significant decrease of |G*| in the mesencephalon, whereas we detected no significant changes of the mesencephalon in the present study. They found no significant effects due to PD on φ (except for the striatum), which mostly agrees with the present findings (see Fig. 5). The comparison of our results and those of Lipp et al^20^ reveals several discrepancies, showing consistently stiffer measurements in the present study, which can be attributed to the use of different methodologies (as described above).

Conventionally, FA and MD (which is qualitatively similar to apparent diffusivity coefficient (ADC)) have been extensively used to study all kinds of neuropathologies, including PD ^43–45^. These studies have provided insights into how white matter tracts are affected by the disease. However, MD and FA are average measures of the entire voxel and yield limited information about the microscopic nature of the investigated tissue. Thus, several studies have also included more advanced dMRI techniques, such as multi-shell dMRI and MD-dMRI to study PD (e.g., References ^8–10,46^) motivated by the fact that measures like μFA (or mean kurtosis-measurements) are more sensitive to microscopic changes than conventional measures like FA and MD^9,46^.

Figure 4 shows how the microstructural properties overall are altered due to aging. A clear reduction of the μFA and an increase of the MD in most structures were found. Furthermore, we report a decrease of the FA for most WM structures, the CC, and an increase of the Var(MD) of the WM of the frontal, temporal, occipital and parietal lobes plus in the Ctx of the temporal lobe. The fact that μFA decreases in the Ctx but not FA can be seen because of the overall low values of FA within the cortex. These results are generally well in line with the results obtained by Refs. ^10,47,48^, who studied the WM of the above-mentioned structures and indeed also detected the same trend, i.e., decreased FA and μFA, and increased MD and Var(MD).

For changes due to PD, Kamiya et al^10^ and Surova et al^8^ also studied the effects of a PD diagnosis on microstructural properties. Kamiya et al.^10^ showed significant effects of the MD in the WM of the parietal, frontal, and occipital lobe (p_uncorr<0.05), whereas we show a similar level of significance only for the temporal and occipital lobes (WM and Ctx) and the mesencephalon. Surova et al.^8^ showed a significant effect with increasing MD for the superior longitudinal fasciculus, a WM tract that connects the temporal, occipital, parietal and frontal lobe, which also qualitatively agrees with the present results.

Kamiya et al^10^ showed very little effect of the μFA (no significant differences between HC and PD in their investigated regions), which is largely in line with what is reported here (although some significant effect was found in the mesencephalon). In the present study, a significant decrease of µFA and FA between HC and PD was found in the mesencephalon and in the temporal WM for the FA signal. This is partly in line with Surova et al^8^, who only found a significant decreasing effect on the putamen and the thalamus, and Kamiya et al^10^ reported some effect on the anterior limb of the internal capsule (p_uncorr<0.05). The largest discrepancy between Kamiya et al.’s study and the present one is that they report several significant effects on the Var(MD) between HC and PD (frontal WM p_uncorr<0.05, parietal WM p<0.05, occipital WM p_uncorr<0.05, temporal WM p_uncorr<0.05) where the size variance increases with PD, whereas in the present study we did not detect any significant differences between HC and PD for Var(MD) in the global regions. However, we found several non-significant (p>0.05) correlations that would agree with Kamiya et al.’s results (Cohen’s d > 0.6 for the mesencephalon, Ctx occipital, WM temporal and occipital).

Several previous studies have investigated the effects of PD on QSM where it is consistently found that QSM levels increase within the substantia nigra (e.g., Refs. ^5,6,49,50^), consistent with the present findings. Acosta-Cabronero et al. (2017) also detected a significant increase of QSM due to PD (after correcting for age effects) within the cortical areas of the lateral occipital, posterior parietal, rostral middle prefrontal, middle temporal gyrus and the hippocampus, which were not apparent in the present study. NMI showed a significant decrease in neuromelanin within the SNc, which is consistent with previous studies^7,51,52^. Changes in both QSM and NMI within the mesencephalon thus agree with what is expected from the literature, which is encouraging for the further use of QSM and NMI as diagnostic markers for PD. It is worth noting that although the mesencephalon in general, and the SN in particular, is usually found to be significantly affected by PD, there were few signs of changes due to mechanical properties in these regions due to PD (see Fig 5b), except a decrease in viscosity due to PD within the VTA. On the other hand, MD-dMRI properties were generally strongly affected, most likely due to neural atrophy (decrease of μFA and FA, increase of MD and Var(MD)). In fact, these modalities were shown to be more significantly affected than QSM and NMI for several regions within the present study.

The decrease in stiffness seen for several regions in the brain due to aging correlates with changes seen in the microstructural parameters obtained from MD-dMRI: reduced stiffness correlates with reduced μFA, and increased MD (also reduced FA in the WM, and increased Var(MD) for some of these regions). In the case of comparing PD to HC (see Fig. 5), these correlations between viscoelastic and MD-dMRI parameters are significantly weaker, which may reveal information about the different microstructural mechanisms behind the effects of aging and PD.

By combining the reported MRE and MD-dMRI-derived parameters, one can better understand the biological mechanisms behind the changes in brain tissue. If we, for example, analyze the WM- regions (which shows very clear signs of softening both due to age and PD effects), we can see that the tissue in these regions decreases in both μFA and FA and increases in MD and Var(MD) due to age. This combination is indicative of neuronal loss due to age: fewer fibers generally results in a lower μFA and a lower FA (assuming we compare to a scenario with generally aligned fibers), which is replaced by extracellular free water, resulting in higher MD and Var(MD) (if the replacement/atrophy occurs heterogeneously). In contrast, these regions appear to be less significantly affected for μFA, FA and Var(MD) when observing effects due to PD. This implies that the neuronal fibers are more intact, or at least that the decrease is too subtle to be significant, whereas the increase of MD implies more free water in the system. This could be explained by, for example, a decrease in the number of non-directional cells (e.g., glial cells) surrounding the axons, which would have a small effect on FA and µFA while allowing more extracellular water in the volume. The cortical gray matter is affected by age- and group-differences in a similar manner to the WM, with the exception that FA remains unchanged for aging effects. This may simply be due to the overall small FA in the cortex, as there is little global anisotropy there, but it should also be noted that there is more noise associated with these regions due to partial volume effects. In the corpus callosum, we note a clear effect of decreasing FA and μFA in combination with increased MD (for aging effects), while little effect is seen on its stiffness. If this motif from the MD-dMRI parameters implies neuronal atrophy or axonal degradation, one would, in contradiction to observed measurements, also presume a loss of stiffness in this region. This discrepancy could possibly be attributed to some form of axonal degradation that does not significantly alter the mechanical properties, such as demyelination. Demyelination is also consistent with an increase in MD without an associated increase in the size variance (Var(MD)), as shown via numerical simulations by Westin et al^34^. Similarly, the mesencephalon exhibit a decrease of µFA and increase of MD due to age, not associated with a change in mechanical properties. These alterations can also be seen for group comparisons, however with the addition of a significant reduction of FA for PD patients, again implying that there is significant axonal degradation (loss of fiber tracts) without any major alterations to the mechanical properties.

Multiple studies have previously shown different effects on the white matter and the cortical grey matter due to PD that fits with the reported findings: Pieperhoff et al^3^, for example, showed that the volume of the temporal and occipital lobe, in particular, is reduced over time for PD patients, indicating that these structures are indeed particularly affected by the disease^‡^. Previous studies using dMRI^8,10^ have also shown some microstructural alterations in the brain due to PD. However, these have been relatively small effects, which fits largely with the presented results. The presented results in this study may give a qualitative indication of how the microstructural mechanisms change due to age and PD and how these couple to the mechanical properties of the tissue. However, further data and analysis are required to support any direct evidence for the discussed mechanisms.

### Limitations

There are several limitations to point out with the presented work. One major limitation of this study is the relatively low number of subjects, which may give rise to statistical uncertainties. This is particularly true for all UPDRS correlations, where we were limited to the PD cohort, consisting of 12 subjects. It should also be stated that UPDRS was performed in ON state and results are a combination of PD *per se* and medication. Another limitation is the relatively low resolution of MRE (3mm) and MD-dMRI (2.5mm), which may give rise to partial volume effects; this is a limitation that will have an effect on the whole brain, where the properties of varying regions spill into each other, but most significantly is this a problem for regions close to CSF where the shear stiffness, for example, is not applicable. Thus, some spurious results may affect the estimations at the cortical surface, for example. This is also true for the analysis of small subcortical structures (e.g., the SNc). The accuracy of the viscoelastic parameters is also limited by the inversion method that relies on common assumptions of the material, such as isotropic viscoelastic properties and local homogeneity^28^.

### Summary and Conclusions

To our knowledge, this is the first study that investigates how the whole brain is affected by PD by combining MRE, MD-dMRI, QSM and NMI. The main findings are:

1. The regions that are most significantly softened by PD are the temporal and the occipital lobes
2. These changes are associated with an increase in the MD, where most other microstructural properties remain insignificantly altered (except a decrease of FA in the WM temporal). This suggests that neural atrophy might not be the predominant mechanism behind the decrease in stiffness.
3. The mesencephalon (and most of its substructures) exhibits alterations of the microstructural properties, much more indicative of neural atrophy. However, these changes are not generally accompanied by changes in the mechanical properties.
4. Multiple regions of the brain are softened due to age, and we show that these changes possibly correlate mainly with a microstructural change that can be attributed to neural atrophy.
5. Mechanical properties are strongly correlated with μFA and MD.

## Acknowledgments

This research has been partially funded by Hälsa, Medicin och Teknik (grant No. 2022-0688), MedTechLabs, Digital Futures, Vinnova through AIDA (project ID: 2319) and the Swedish Research Council (grant No. 2022-03389). The funding sources were not involved in the research and preparation of this article. We would like to thank the participants in this study and the clinicial research scientists at Philips and MR physicists at Karolinska University hospital in Huddinge, who were involved in setting up the imaging protocols and for their help with postprocessing. We would also like to thank Prof. Richard Ehman and his group at Mayo clinic for providing the MRE capability and for their support with this, and Marcus Nilsson and his group at Lund University for helping providing and supporting the MD-dMRI pipeline.

## Author contributions

CO: Conceptualization, Methodology, Software, Formal analysis, Investigation, Visualisation, Writing – original draft, Writing – review and editing. MS: Conceptualization, Methodology, Writing – review and editing. PS: Conceptualization, Methodology, Funding acquisition, Writing – review and editing. RM: Conceptualization, Methodology, Supervision, Resources, Funding acquisition, Writing – review and editing, Project administration.

## Data availability

The MR imaging data in this study are subject to ethical and legal restrictions and cannot be shared publicly, however anonymized data can be made available upon reasonable request to researchers who meet the legal requirements. Please contact the corresponding author for more information.

## Footnotes

* The FOV was occasionally, for some subjects, cut off at the inferior part of the cerebellum, which is why this part of the anatomy is omitted from any analysis.

† Both studies included a cohort of patients with progressive supranuclear palsy (PSP), which are excluded from any comparisons presented here.

‡ They also showed an effect in the inferior parietal lobule, the insula, putamen and the nucleus basalis of Meynert

