## Supplementary Information for "Effects of Parkinson’s Disease on Mechanical and Microstructural Properties of the Brain"

#### 1. Comparisons to previous studies

*MRE results compared to results from Lipp et al.*

In Table SI1, we compare the effect of age on the stiffness. In addition to the whole brain stiffness, we also report the stiffness of the lentiform nucleus in order to compare them with the results from Lipp et al. (2018, 2013). Compared to these studies, we find a stronger effect of age on the whole brain on both HC and PD, possibly due to a wider age range. We furthermore find a less significant impact of age on the lentiform nucleus than the one reported by Lipp et al. (2013).

Table SI1. Correlations between stiffness and age, Pearson r-values (p-value) and significant values are shown in bold. Values from other studies are Lipp et al. (2018, 2013)[1,2]

| | Whole brain, HC | Lentiform Nucleus HC | Whole brain, PD | Lentiform Nucleus PD |
|---|---|---|---|---|
| Corr(\|G*\|age) (p-value). Present study | **-0.6** (0.0083) | -0.45 (0.07) | **-0.91** (0.00012) | -0.41 (0.21) |
| Corr(\|G*\|age) (p-value) Lipp et al. (2013) | **-0.47** (0.048) | N/A | **-0.49** (<0.05) | **-0.76**(p<0.05) |
| Corr(\|G*\|age) (p-value) Lipp et al. (2018) | 0.2 (0.54) | N/A | **-0.47** (0.06) | N/A |

Table SI2 shows the average stiffness and viscosity of the whole brain (excluding regions containing CSF), the lentiform nucleus and the frontal region both for our cohort and the cohorts from Lipp et al. (2013, 2018). The obtained stiffness values in the brain from this study were comparable to those reported in Lipp et al. (2013), using their 3DMRE method, despite the fact that the methods of obtaining their measurements differed from the ones used here. However, the reported stiffnesses by Lipp et al. (2018) are significantly lower than those in their previous study and our presented measurements. This discrepancy could possibly be due to the fact that they used multifrequency acquisition and a different postprocessing pipeline in Lipp et al. (2018).

*Table SI2. Stiffness and viscosity values compared with previous PD MRE studies. Stiffness, |G*|, values are given in Pa.*

| | Whole brain, HC | Whole brain, PD | Lentiform Nucleus HC | Lentiform Nucleus PD | Frontal HC | Frontal PD |
|---|---|---|---|---|---|---|
| \|G*\| (present study) | 2034±148 | 1894±174 | 2414±248 | 2290±293 | 2219±259 | 2040±306 |
| φ (present study) | 0.34±0.02 | 0.32±0.02 | 0.37±0.07 | 0.34±0.09 | 0.37±0.05 | 0.32±0.11 |
| \|G*\| Lipp 2013 (3DMRE) | 1970±176 | 1876±255 | 2101±199 | 1955±213 | - | - |
| φ Lipp 2013 (3DMRE) | 0.26±0.04 | 0.22±0.07 | 0.29±0.08 | 0.23±0.11 | - | - |

| | | | | | | |
|---|---|---|---|---|---|---|
| \|G*\| Lipp 2018 (3DMRE) | 1040±80 | 960±65 | - | - | 1150±214 | 990±169 |
| φ Lipp 2018 (3DMRE) | 0.54±0.05 | 0.51±0.05 | - | - | 0.59±0.06 | 0.57±0.06 |

#### 2. Right and left asymmetries

***Age effects after group correction***

Changes in stiffness due to age are relatively symmetric. The viscosity of the right hemisphere (rh) temporal WM is more affected than the left hemisphere (lh), and there is a slight difference in viscosity between parietal Ctx. lh WM temporal is more affected than rh in μFA, and rh occipital Ctx reduces less in μFA than its left side. Left WM frontal, parietal, and temporal increase more in Var(MD) with age than the right side.

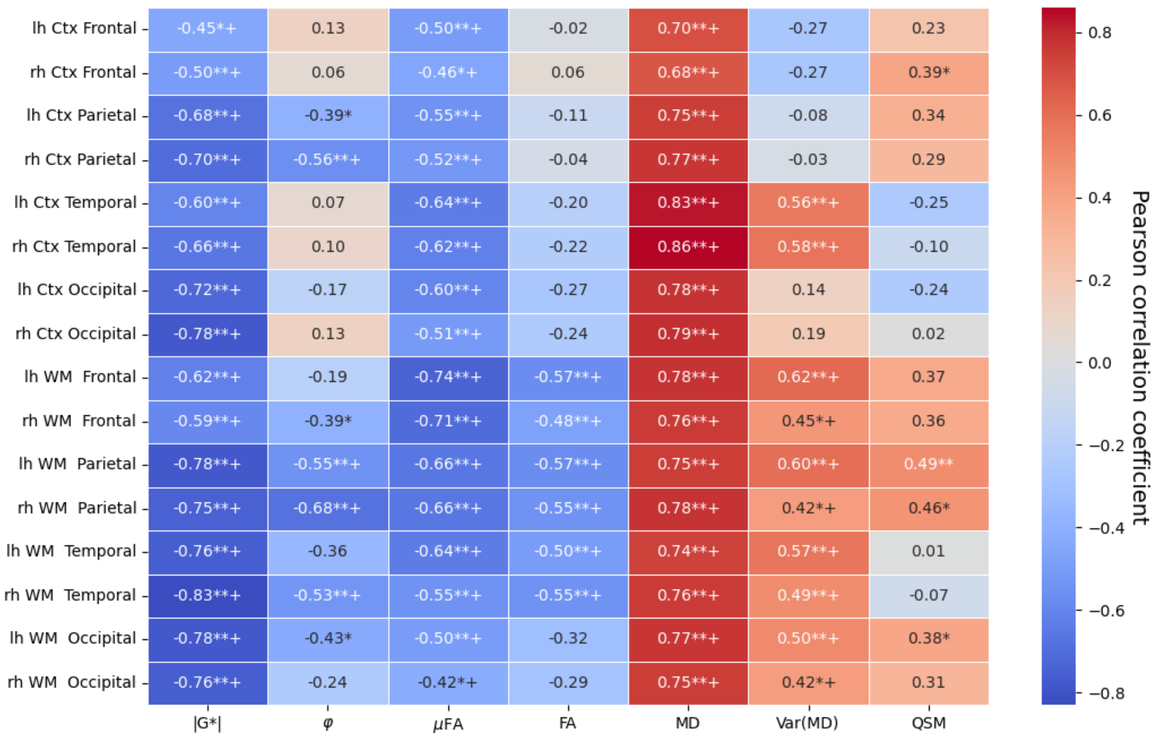


*Figure SI1. Correlations with age after group (HC:PD) correlations for cerebral regions separated by left hemisphere (lh) and right hemisphere (rh). Values shown are Pearson correlations, where *=p<0.05, **=p<0.01, +=p_FDR-corr<0.05*

***Group effects:***

Stiffness of the left side reduces more than in the rh due to PD for parietal and occipital Ctx, and for the occipital WM. No significant differences between rh and lh were seen for the other modalities.

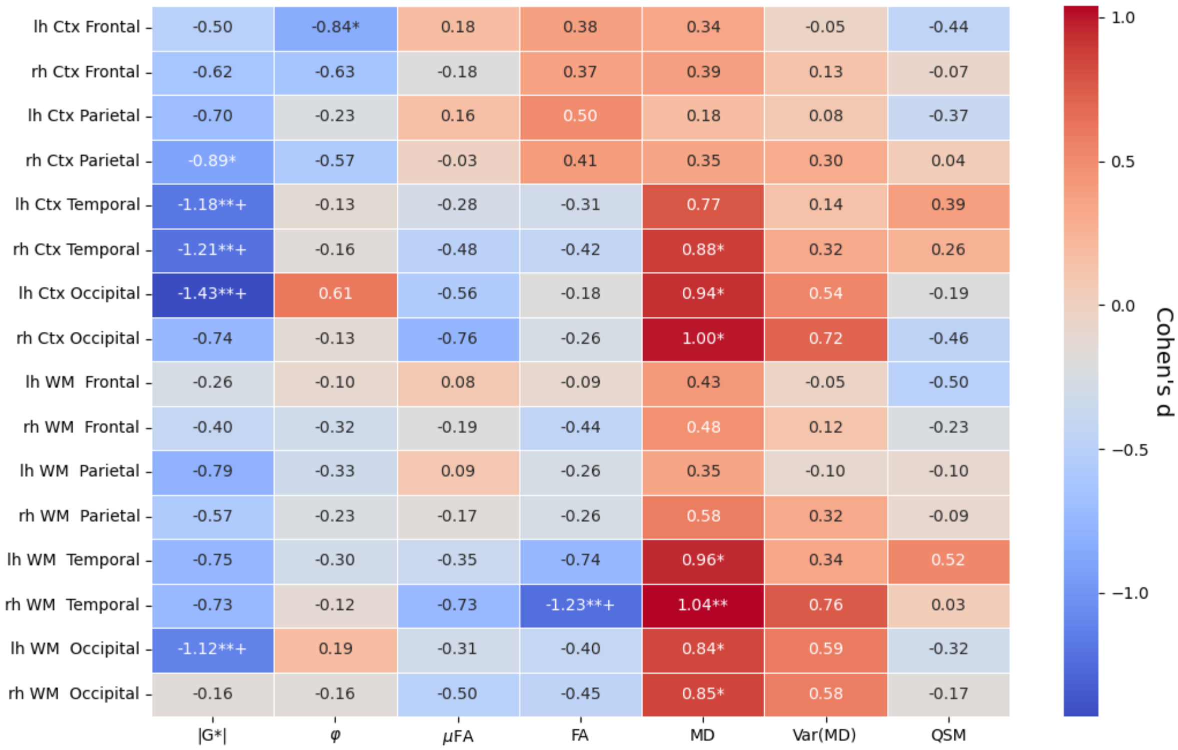


*Figure SI2. PD effects after age correlations for cerebral regions separated by left hemisphere (lh) and right hemisphere (rh). Correlation strengths are Cohen's distances between the two groups, and p-values are calculated based on Mann-Whitney U-test between the groups. . *=p<0.05, **=p<0.01, +=p_FDR-corr<0.05*

#### 3. UPDRS scores

Here, we calculate how the different modalities correlate (using Pearson correlation as a correlation measure, with age as a covariate) with various UPDRS scores from the patient cohort. We show the subcortical structures specifically due to their typical importance related to PD, and because we do not capture the global values of NMI since the FOV was focused on the midbrain area for this modality.

##### UPDRS 1:

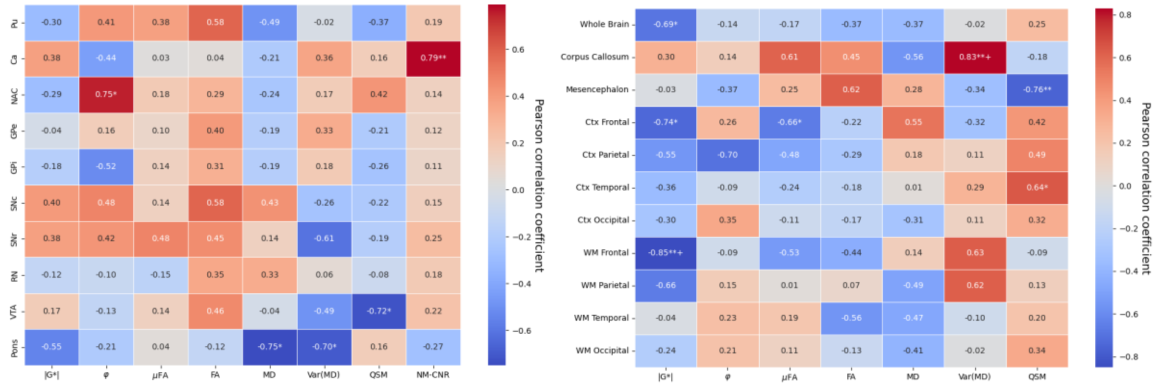


*Figure SI3. Correlations between changes in the measured values with UPDRS 1 score (age-corrected) for the PD cohort (excluding HC). Left) Subcortical structures Right) Global structures *=p<0.05, **=p<0.01, +=p_FDR-corr<0.05*

### UPDRS 2

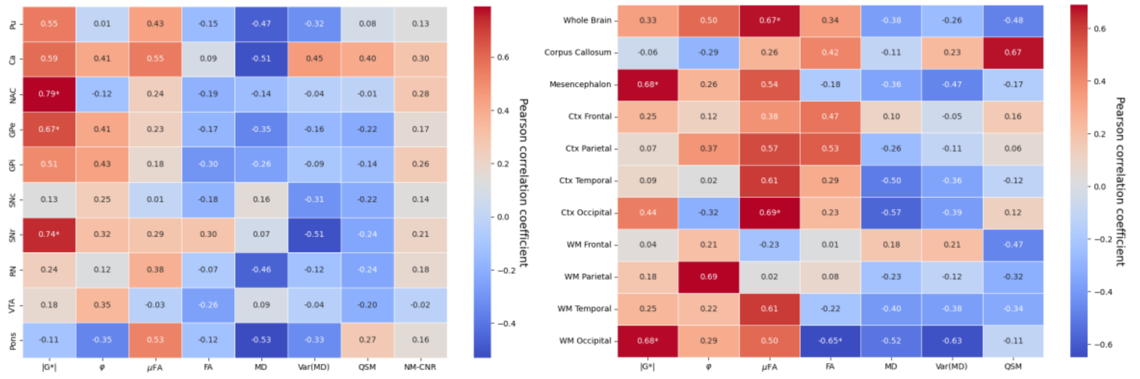


*Figure SI4 Correlations between changes in the measured values with UPDRS 2 score (age-corrected) for the PD cohort (excluding HC). Left) Subcortical structures Right) Global structures *=p<0.05, **=p<0.01, +=p_FDR-corr<0.05*

### UPDRS 3

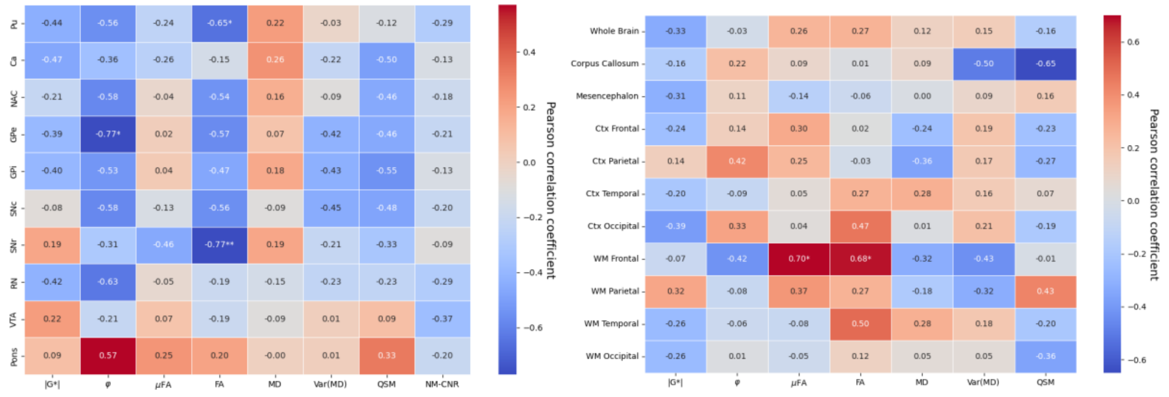


*Figure SI5 Correlations between changes in the measured values with UPDRS 3 score (age-corrected) for the PD cohort (excluding HC). Left) Subcortical structures Right) Global structures *=p<0.05, **=p<0.01, +=p_FDR-corr<0.05*

### UPDRS 4:

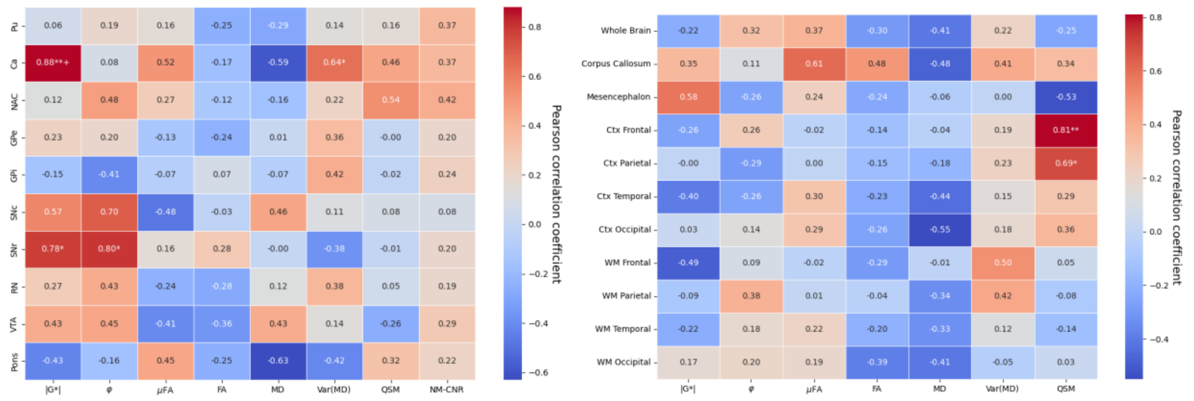


*Figure SI5. Correlations between changes in the measured values with UPDRS 4 score (age-corrected) for the PD cohort (excluding HC). Left) Subcortical structures Right) Global structures. *=p<0.05, **=p<0.01, +=p_FDR-corr<0.05*

### 4. Group comparisons

***Brain regions***

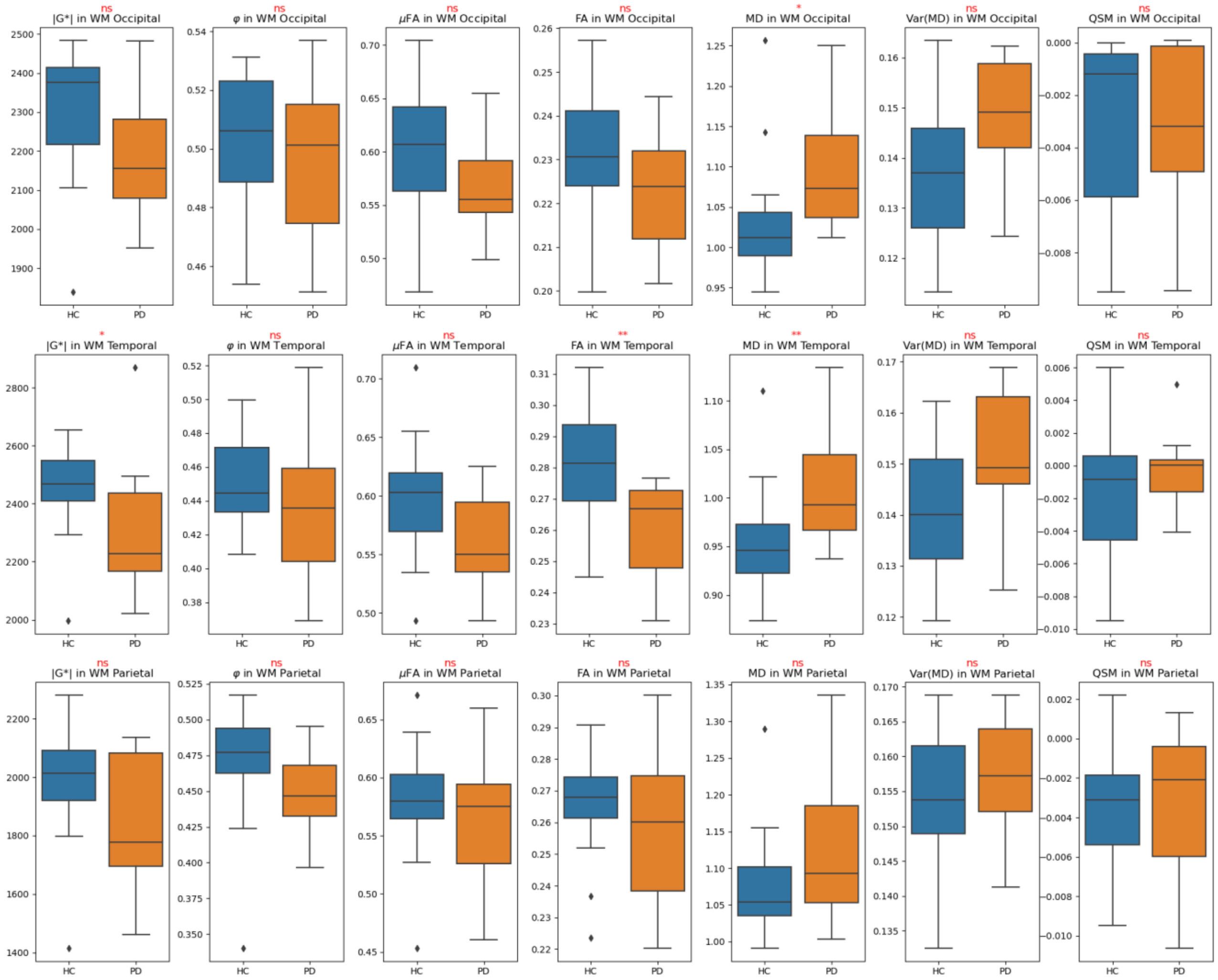

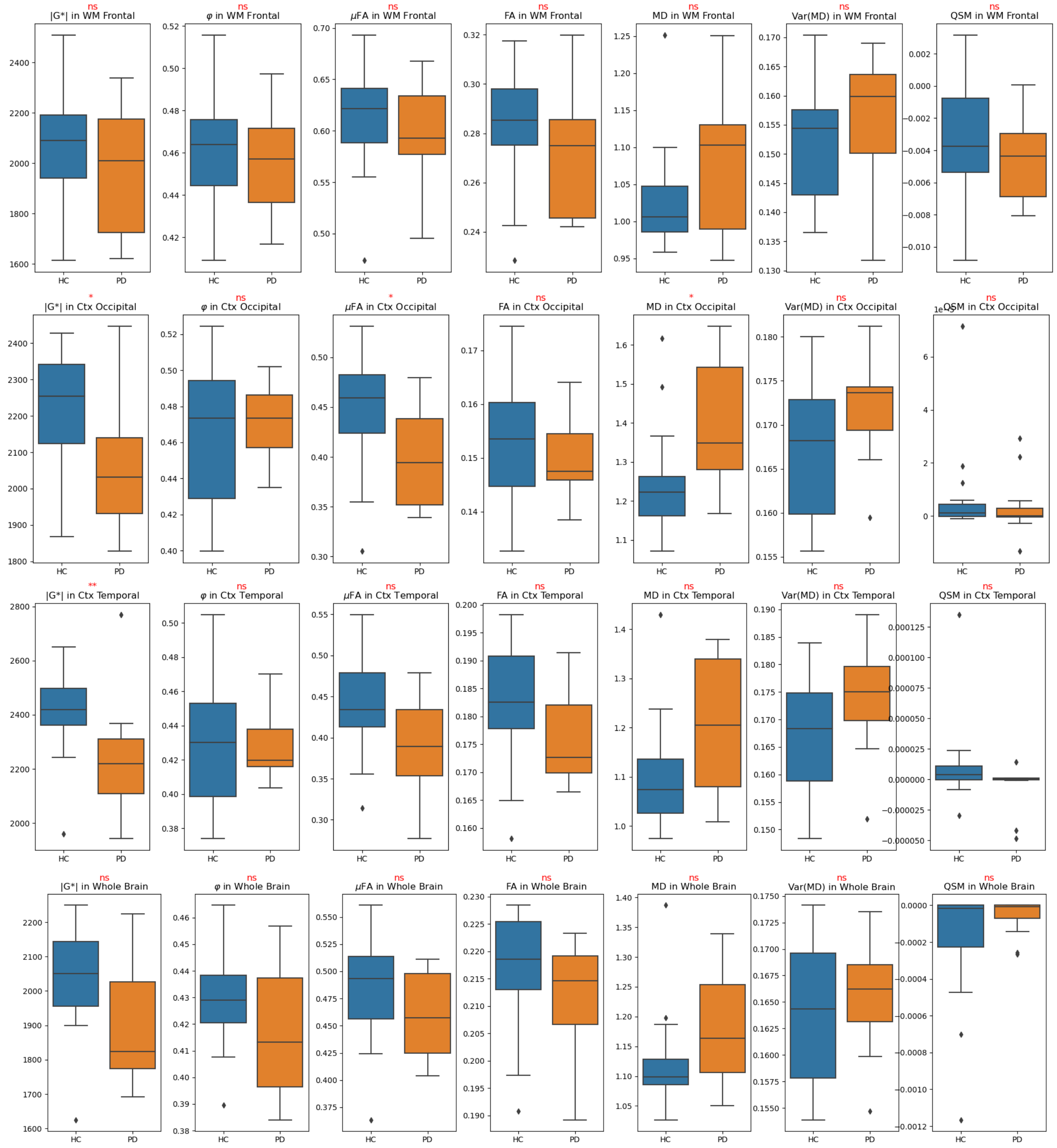
ns |G*| in WM Frontal
ns φ in WM Frontal
ns μFA in WM Frontal
ns FA in WM Frontal
ns MD in WM Frontal
ns Var(MD) in WM Frontal
ns QSM in WM Frontal
* |G*| in Ctx Occipital
ns φ in Ctx Occipital
* μFA in Ctx Occipital
ns FA in Ctx Occipital
* MD in Ctx Occipital
ns Var(MD) in Ctx Occipital
ns QSM in Ctx Occipital
1e-5
** |G*| in Ctx Temporal
ns φ in Ctx Temporal
ns μFA in Ctx Temporal
ns FA in Ctx Temporal
ns MD in Ctx Temporal
ns Var(MD) in Ctx Temporal
ns QSM in Ctx Temporal
ns |G*| in Whole Brain
ns φ in Whole Brain
ns μFA in Whole Brain
ns FA in Whole Brain
ns MD in Whole Brain
ns Var(MD) in Whole Brain
ns QSM in Whole Brain
HC
PD

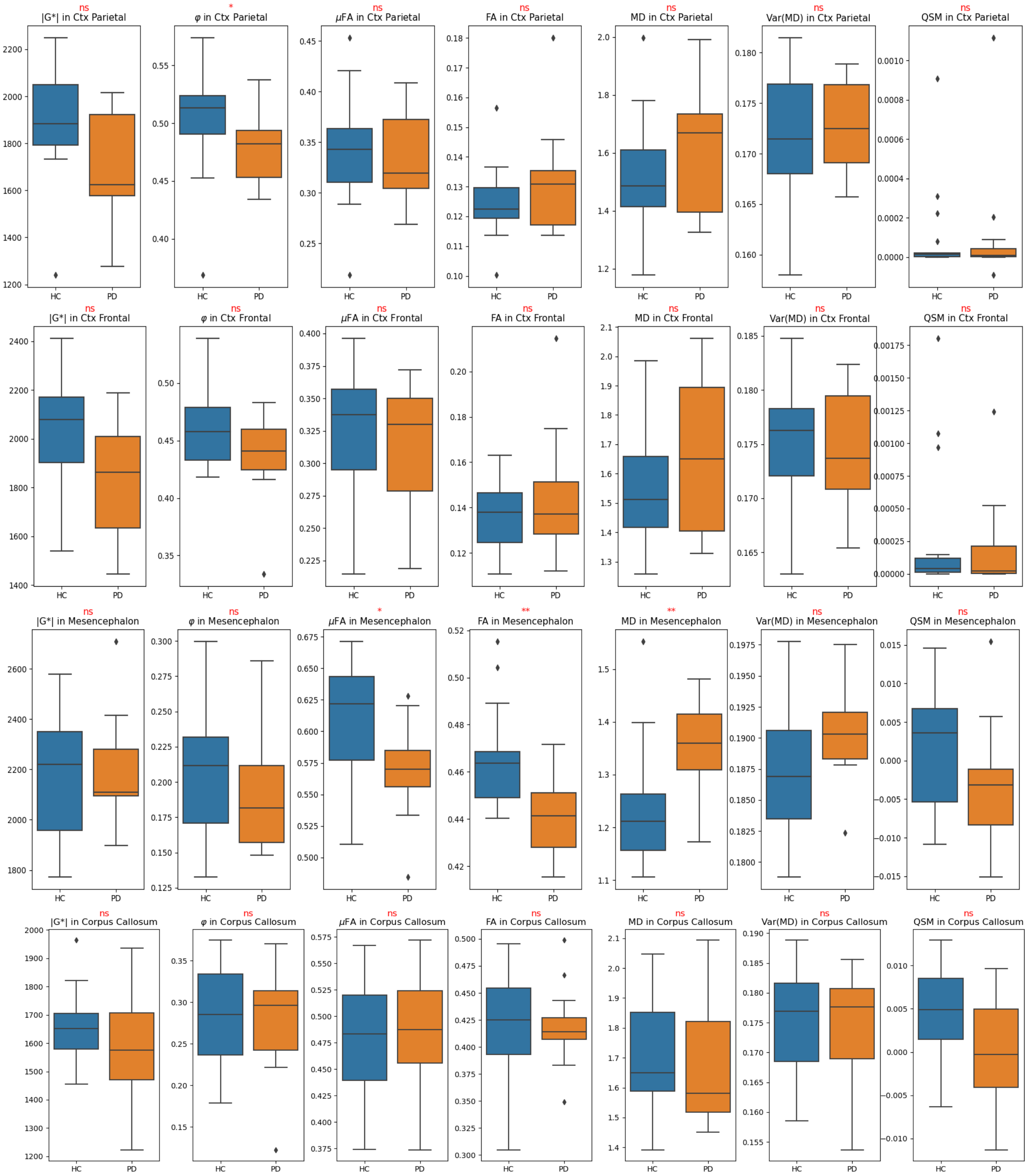


*Figure SI6. Boxplots showing the median value in all global regions for all imaging modalities. Blue boxplots show all healthy controls (HC) and orange boxplots show data for all PD patients. Image modality and region are displayed in the title for each subplot, where we also display the level of significance of the difference between the two groups (ns=no significance/p>0.05, *=p<0.05, **p<0.01, no FDR correction applied).*

***Subcortical structures***

Figure SI7. Boxplots showing the median value in all subcortical regions for all imaging modalities. Blue boxplots show all healthy controls (HC) and orange boxplots show data for all PD patients. Image modality and region are displayed in the title for each subplot, where we also display the level of significance of the difference between the two groups (ns=no significance/p>0.05, *=p<0.05, **p<0.01, no FDR correction applied).

#### 5. Correlations among image modalities

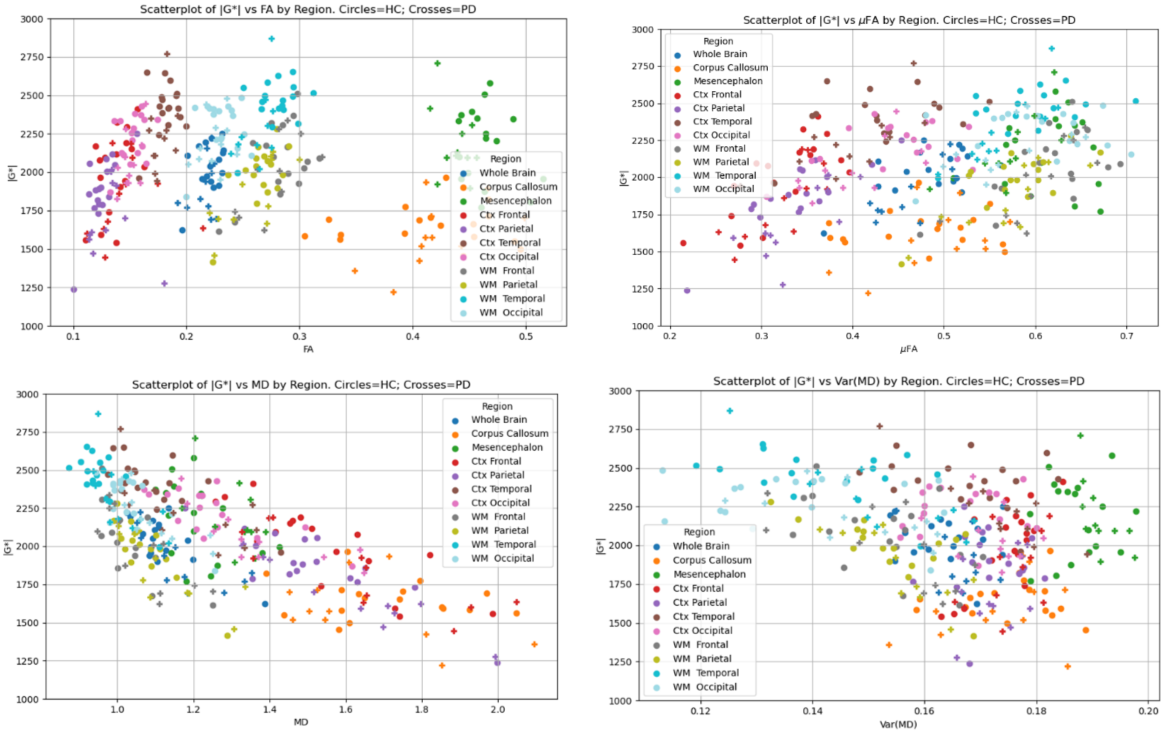


Figure SI 8 |G*| (in Pa) as a function of various MD-dMRI parameters. For all subjects divided into different regions as listed in the legend. Circles and crosses are used to mark HC and PD subjects, respectively.

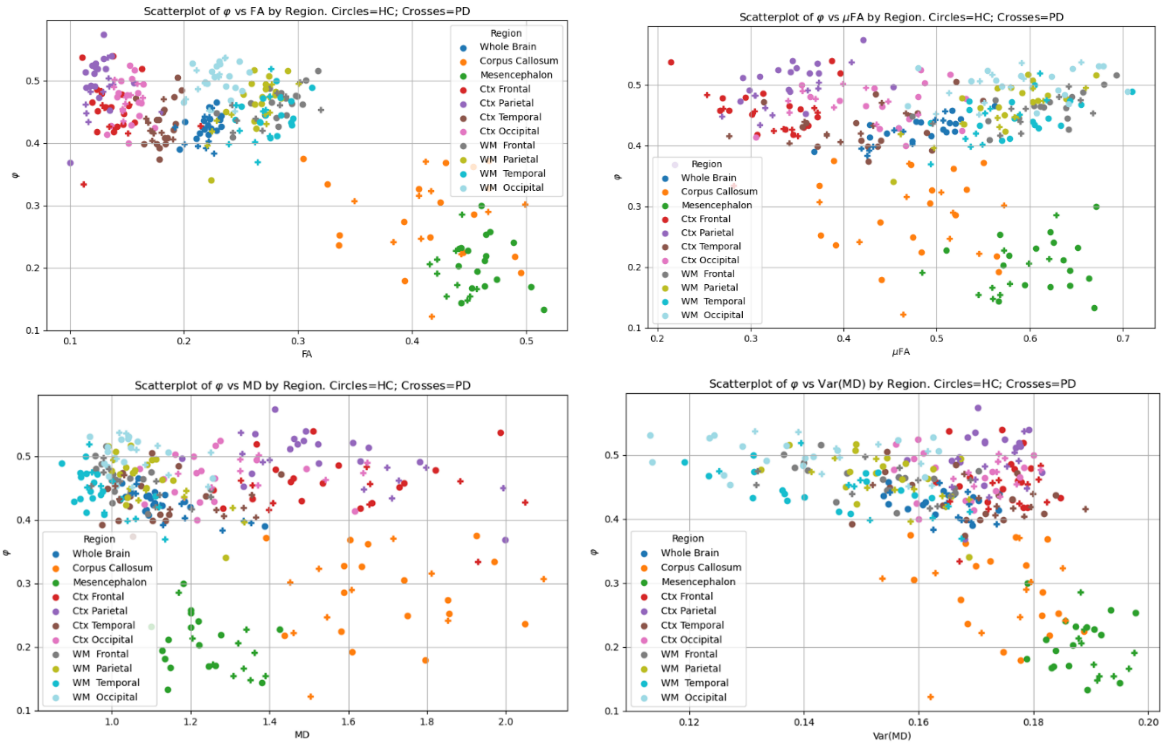


Figure SI 9 Viscosity angle (φ) in radians, as a function of various MD-dMRI parameters. For all subjects divided into different regions as listed in the legend. Circles and crosses are used to mark HC and PD subjects, respectively.

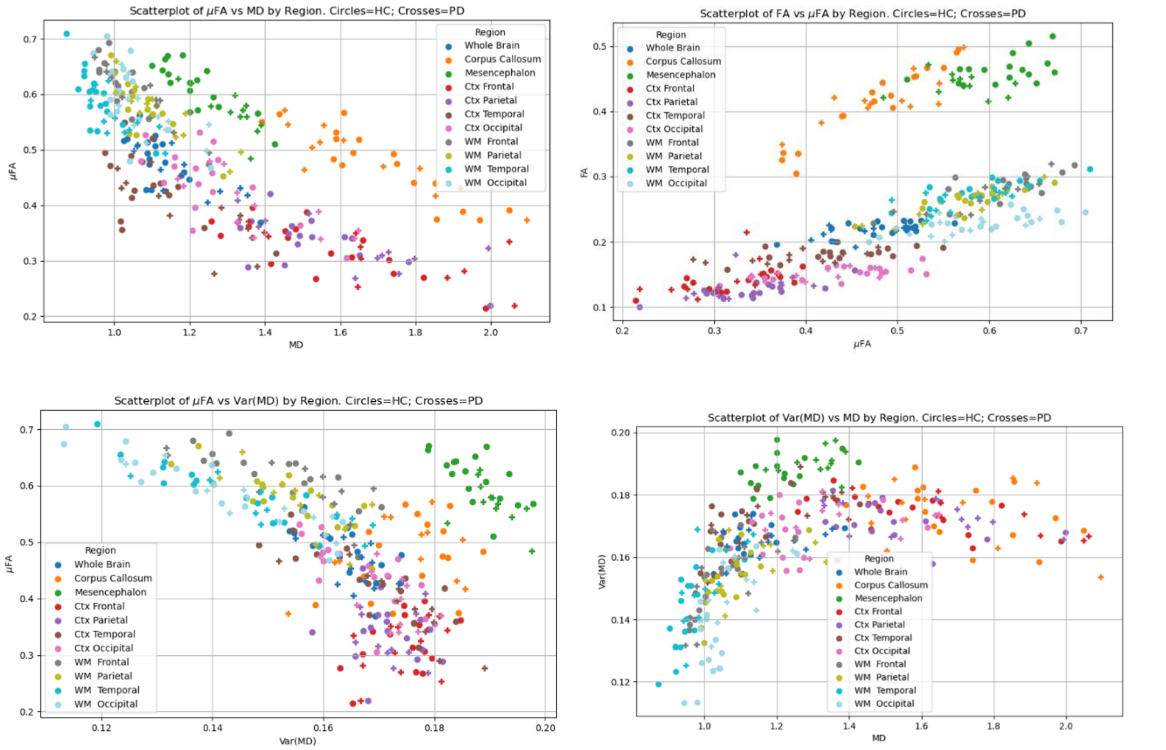

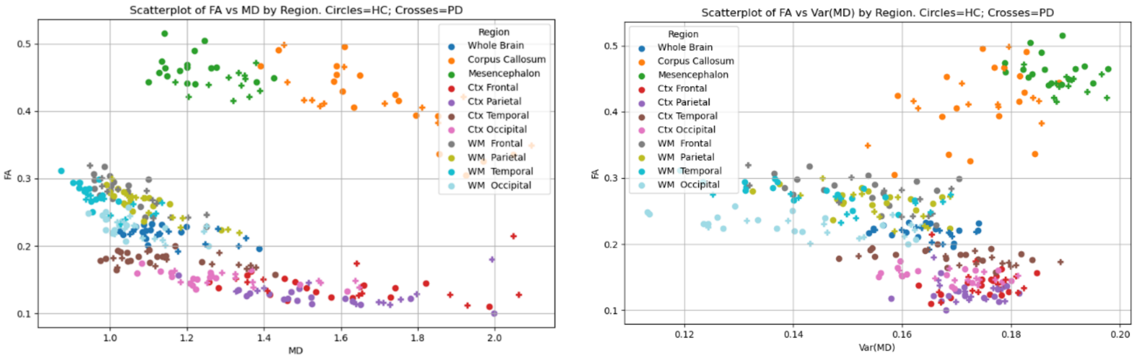


Figure SI 10 Correlations between various MD-dMRI parameters. For all subjects divided into different regions as listed in the legend. Circles and crosses are used to mark HC and PD subjects, respectively.

#### References SI

1. Lipp, A. *et al.* Progressive supranuclear palsy and idiopathic Parkinson's disease are associated with local reduction of in vivo brain viscoelasticity. *Eur. Radiol.* **28**, 3347–3354 (2018).

2. Lipp, A. *et al.* Cerebral magnetic resonance elastography in supranuclear palsy and idiopathic Parkinson's disease. *NeuroImage Clin.* **3**, 381–387 (2013).
